# Anti-Diabetic and Antiresorptive Pharmacotherapies for Prevention and Treatment of Type 2 Diabetes-Induced Bone Disease: Protocol for a Two-Part Systematic Review and Network Meta-Analysis

**DOI:** 10.1101/19007849

**Authors:** Jiawen Deng, Umaima Abbas, Oswin Chang, Sayan Dhivagaran, Stephanie Sanger, Anthony Bozzo

## Abstract

**Introduction:** Patients with type 2 diabetes mellitus (T2DM) are at risk for a variety of severe debilitating effects. One of the most serious complications experienced by T2DM patients are skeletal diseases caused by changes in the bone microenvironment. As a result, T2DM patients are at risk for higher prevalence of fragility fractures.

There are a variety of treatments available for counteracting this effect. Some anti-diabetic medications, such as metformin, have been shown to have a positive effect on bone health without the addition of additional drugs into patients’ treatment plans. Chinese randomized controlled trial (RCT) studies have also proposed antiresorptive pharmacotherapies as a viable alternative treatment strategy. Previous network meta-analyses (NMAs) and meta-analyses regarding this topic did not include all available RCT trials, or only performed pairwise comparisons. We present a protocol for a two-part NMA that incorporates all available RCT data to provide the most comprehensive ranking of anti-diabetics (Part I) and antiresorptive (Part II) pharmacotherapies in terms of their ability to decrease fracture incidences, increase bone mineral density (BMD), improve indications of bone turnover markers (BTMs), and decrease pain in adult T2DM patients.

**Methods and Analysis:** We will search MEDLINE, EMBASE, PubMed, Web of Science, CINAHL, CENTRAL and Chinese literature sources (CNKI, CQVIP, Wanfang Data, Wanfang Med Online) for randomized controlled trials (RCTs) which fit our criteria. We will include adult T2DM patients who have taken anti-diabetics (Part I) or antiresorptive (Part II) therapies with relevant outcome measures in our study.

We will perform title/abstract and full-text screening as well as data extraction in duplicate. Risk of bias (RoB) will be evaluated in duplicate for each study, and the quality of evidence will be examined using CINeMA in accordance to the GRADE framework. We will use R and gemtc to perform the NMA. We will report changes in BMD, BTM and pain scores in either weighted or standardized mean difference, and we will report fracture incidences as odds ratios. We will use the surface under the cumulative ranking curve (SUCRA) scores to provide numerical estimates of the rankings of interventions.

**Ethics and Dissemination:** The study will not require ethics approval. The findings of the two-part NMA will be disseminated in peer-reviewed journals and presented at conferences. We aim to produce the most comprehensive quantitative analysis regarding the management of T2DM bone disease. Our analysis should be able to provide physicians and patients with up-to-date recommendations for anti-diabetic medications and antiresorptive pharmacotherapies for maintaining bone health in T2DM patients.

**Systematic Review Registration:** International Prospective Register for Systematic Reviews (PROSPERO) — CRD42019139320

**ARTICLE SUMMARY:** *Strengths and limitations of this study:*

- Literature search in Chinese databases will yield new RCT evidence regarding the efficacy of anti-diabetics in treating T2DM bone disease
- Using network meta-analytical techniques to analyze the relative efficacy of antiresorptive therapies will allow us to include new treatment arms, such as zoledronic acid and risedronate.
- Only RCTs will be included and the quality of trials and networks will be evaluated using Risk of Bias, GRADE and comparison-adjusted funnel plots.
- Chinese clinicians may not use the same procedures and practices as Western clinicians, therefore the outcomes from Chinese RCTs may not apply to the Western healthcare systems.
- The study design does not allow the comparison of anti-diabetics with antiresorptive therapies or combinations of the two.

## Introduction

Diabetes mellitus (DM) is an epidemic collection of metabolic diseases featuring substantial morbidity and mortality around the globe. Type 2 diabetes mellitus (T2DM), which constitutes 90-95% of all adult DM cases in the US, is the most common type of DM[1]. T2DM is characterized by relative insulin deficiency, stemming from pancreatic β-cell dysfunction and insulin resistance in organs[2]. T2DM can be caused by a variety of factors, including excess body weight, physical inactivity, as well as sugar and fat consumption[3]. Over the past decades, there has been a significant increase in the incidence of T2DM around the world, from 108 million in 1980 to 451 million in 2017[4,5]. As a result of this trend, the number of people with T2DM globally is expected to increase to 693 million by 2045[5]. With rising incidence, it is crucial for physicians to be informed of the most optimal clinical strategies to counteract T2DM’s debilitating effects.

One of the many complications that T2DM patients suffer from are skeletal weakness and fragility fractures[6]. Patients with T2DM experience accelerated bone resorption, impaired osteoblast-mediated bone formation, and poorer bone quality compared to those without T2DM[7]. Research shows that hyperglycemia as a result of insulin resistance can lead to the production of advanced-glycation end-products (AGEs) in collagen, which stimulate apoptosis of osteoblasts and induce abnormal arrangement and alignment of collagen[8]. The effect of AGEs on the bone microenvironment, along with abnormal cytokine production and impaired neuroskeletal functions, put T2DM patients at a higher risk for skeletal conditions such as osteoporosis and Charcot’s arthropathy[9,10].

There have been several large-scale observational studies investigating associations between bone mineral density (BMD) — an indicator for osteoporosis and a surrogate marker for fragility fractures — and T2DM. These studies all produced contradictory results with higher, lower, or similar values when compared to healthy controls. The inconsistencies were likely due to differences in their study methodologies, including differences in sites of BMD measurement and bone densitometry techniques, as well as the demographics of the study population[11–14]. However, previous studies have demonstrated that T2DM patients experience an increased risk of fractures independent of BMD[15–17]. Bone turnover markers (BTMs), which is an indicator for the rate of bone formation and resorption, has been shown to deteriorate in T2DM patients as well[18]. These signs and symptoms, combined with high prevalence of vertebral bone pain in the T2DM population, suggest that managing T2DM-induced bone disease is crucial to improving the patients’ quality of life and clinical outcomes[19].

Recent studies have shown that some anti-diabetic medications, namely metformin and sulfonylureas, have a positive effect on bone health and may potentially lower fracture incidence in T2DM patients[20,21]. Hence, anti-diabetic medications can be used as a potential treatment strategy for T2DM bone disease without having to introduce new medications into patients’ treatment plans. However, this effect is not observed in every class of anti-diabetics. Sodium-glucose co-transporter 2 (SGLT2) inhibitors, for example, can increase bone resorption and negatively affect bone health in T2DM patients[22]. Meanwhile, a series of large scale randomized controlled trials (RCTs) have presented alternative strategies to combating T2DM bone disease by using antiresorptive therapies — such as bisphosphonates, calcitonin, vitamin D and calcium supplementations — with promising results[23,24].

We identified two previous network meta-analyses (NMAs) that evaluated the impact of anti-diabetic medications on fracture risks in T2DM patients; however, these studies focused only on SGLT2 inhibitors and thus did not incorporate all available RCT data[25,26]. We identified a single meta-analysis from China regarding the use of alendronate as an antiresorptive therapy in T2DM patients; nonetheless the meta-analysis did not account for all available antiresorptive treatment arms[27].

Therefore, we propose to conduct a two-part systematic review and NMA of RCTs to investigate the following research questions: What are the comparative effects (in terms of fracture incidences) of different anti-diabetic and antiresorptive pharmacotherapies on adult T2DM patients? We will also investigate the comparative effects of these drugs on BMD, BTMs, and bone pain as our secondary outcomes. We will compare anti-diabetic medications for Part I of our analysis, and antiresorptive pharmacotherapies for Part II of our analysis.

## METHODS AND ANALYSIS

We will conduct this two-part systematic review and NMA in accordance to the Preferred Reporting Items for Systematic Reviews and Meta-Analyses (PRISMA) incorporating NMA of health care interventions[28]. This study is prospectively registered on The International Prospective Register of Systematic Reviews (PROSPERO) — CRD42019139320. Any significant amendments to this protocol will be reported and published with the results of the review.

### Eligibility Criteria

#### Types of Participants

We will include adult patients (18 years or older) who have been diagnosed with T2DM according to criteria recommended by the World Health Organization (WHO), the American Diabetes Association (ADA), or the International Diabetes Federation (IDF)[29–31].

Our database search will likely produce studies with a broad range of publication dates; consequently, we may see different sets of criteria from WHO, ADA and IDF as these recommendations tend to be updated periodically. To include a sufficient amount of data for analysis, we will not place restrictions regarding the exact set of criteria used by the study.

Patients included in Part I of the analysis should not receive any form of additional antiresorptive therapies that can affect bone metabolism. However, because anti-diabetic medications are sometimes crucial for stopping the progression of T2DM, anti-diabetic therapies will be allowed for Part II of the analysis due to ethical concerns.

Patients labelled as “pre-diabetic” as defined by the diagnostic criteria will not be included for this study.

#### Types of Studies

We will include parallel-groups RCTs. If a RCT uses a crossover design, latest data from before the first crossover will be used.

#### Types of Interventions

We will include any commonly used anti-diabetic medications for Part I of the analysis. This may include (but not limited to) sulfonylureas, meglitinides, biguanides, thiazolidinediones, alpha-glucosidases inhibitor, dipeptidyl-peptidase-4 (DPP-4) inhibitors, glucagon-like peptide-1 (GLP-1) agonist, and sodium glucose cotransporter 2 (SGLT2) inhibitors. If data permits, placebo, insulin supplementation, and/or lifestyle changes/no pharmacotherapy treatment will also be included. Because concurrent therapies are common in clinical settings, any combinations of anti-diabetic therapies will be included as treatment arms as well.

We will include any antiresorptive pharmacotherapies used to manage bone loss for Part II of the analysis. This may include (but not limited to) bisphosphonates (e.g. alendronate, risedronate, zoledronic acid), calcitonin, calcium, vitamin D or D analogs (e.g. calcitriol or alfacalcidol). If data permits, placebo and untreated (i.e. no antiresorptive treatment) will also be included as treatment arms. We will include combinations of multiple antiresorptive therapies.

We will differentiate treatment arms by daily dosages (e.g. alendronate 5 mg v. alendronate 10 mg); however, if there are RCTs that cannot be included into the network due to the inclusion of dosages, we will disregard dosages and combine treatment arms to facilitate network connections.

### Primary Outcomes

#### Fracture Incidence

We will evaluate fracture incidences based on data collected at the latest follow-up. If data permits, we will conduct separate analyses for vertebral and nonvertebral fractures. Definitions of fractures will be defined as per individual study criteria.

### Secondary Outcomes

#### Change in BMD

We will evaluate change in BMD from baseline, in both percentage and absolute change. BMD change must be calculated based on BMD data collected at the latest follow-up.

We will analyze BMD readings taken at the lumbar spine, femoral neck, total hip, Ward’s Triangle and the greater trochanter. Absolute and percentage changes in T-score and Z-score will not be included in this analysis.

#### Change in Bone Turnover Markers

We will analyze the following BTMs in our NMA:

- Bone resorption biomarkers: Tartrate-Resistant Acid Phosphatase 5b (TRAP 5b), Carboxy-terminal Crosslinked Telopeptide of type 1 collagen (CTX-1), Amino-terminal Crosslinked Telopeptide of type 1 collagen (NTX-1).
- Bone formation biomarkers: Bone Alkaline Phosphatase (BAP), Osteocalcin (OC), Procollagen type 1 N-terminal Propeptide (P1NP).

These BTMs are chosen for their common use in the investigation of bone diseases and the availability of extensive literature regarding their applications[32,33]. While our preliminary database search has shown that there are several large scale RCTs that reported some of these BTMs, the availability of BTM data in our target literature sources was not a factor in our method design[34].

Change in BTM levels will be recorded as percentage changes from baseline. We will include only percentage changes calculated using the BTM level measured at the latest follow-up in our analysis.

#### Change in Bone Pain Score

We will include absolute change in bone pain score from baseline using final values measured during the latest follow-up period. We will include pain score measured using any pain scale in the analysis.

### Search Methods for Identification of Studies

#### Electronic Database Search

We will conduct a librarian-assisted search of Medical Literature Analysis and Retrieval System Online (MEDLINE), Excerpta Medica Database (EMBASE), Web of Science, Cumulative Index to Nursing and Allied Health Literature (CINAHL), and Cochrane Central Register of Controlled Trials (CENTRAL) from inception to October 2019. We will use relevant Medical Subject Headings (MeSH) terms to ensure broad and appropriate inclusions of titles and abstracts (see **Supplementary Data**).

Major Chinese databases, including Wanfang Data, Wanfang Med Online, China National Knowledge Infrastructure (CNKI), and Chongqing VIP Information (CQVIP) will also be searched using a custom Chinese search strategy (see **Supplementary Data**).

A single, comprehensive set of search strategies will be used to identify studies relevant to both parts of the analysis. We will not perform separate database searches for both parts of the analysis.

#### Other Data Sources

We will hand search the reference list of previous meta-analyses and NMAs for included articles. We will also review clinicaltrials.gov and WHO International Clinical Trials Registry Platform (WHO-ICTRP) for registered published or unpublished studies.

### Data Collection and Analysis

#### Study Selection

We will perform title and abstract screening independently and in duplicate using Rayyan QCRI[35]. Studies will only be selected for full-text screening if both reviewers deem the study relevant, to either Part I or Part II of the analysis.

Full-text screening will also be conducted in duplicate. We will resolve any conflicts via discussion and consensus or by recruiting a third author for arbitration. We will identify articles specific to Part I and II and separate them at this stage of article screening. Due to our inclusion criteria, we do not expect any article to be included in both Part I and II.

#### Data Collection

We will carry out data collection independently and in duplicate using data extraction sheets developed *a priori*. We will resolve discrepancies by recruiting a third author to review the data. The extraction sheets are similar for both parts of the analysis, as described in the **Data Items** section.

#### Risk of Bias

We will assess risk of bias (RoB) independently and in duplicate using The Cochrane Collaboration’s tool for assessing risk of bias in randomized trials[36]. Two reviewers will assess biases within each article in seven domains: random sequence generation, allocation concealment, blinding of participants and personnel, blinding of outcome assessment, incomplete outcome data, selective reporting, and other sources of bias.

If a majority of domains are considered to be low risk, the study will be assigned a low RoB. Similarly, if a majority of domains are considered to be high risk, the study will be assigned a high RoB. If more than half of the domains have unclear risk or if there is a balance of low and high risk domains, the study will be assigned an unclear RoB.

#### Special Considerations for Chinese Trials

Chinese RCTs are often reported with a poor description of blinding, randomization, and allocation concealment techniques. This is partially due to Chinese clinicians’ inadequate understanding of RCT designs; we also speculate that limitations in the format of Chinese journal articles, which are often restricted to shorter lengths (1-2 pages) compared to Western studies, forced Chinese authors to condense descriptions of their methodology[37].

Because of these factors, we will report RoB results separately for Western and Chinese articles. If we observe significant differences in RoB between the two sets of articles, we will include additional analyses in the supplementary material of the final publication(s) with Chinese and English RCTs being analyzed separately.

### Data Items

#### Bibliometric Data

Authors, year of publication, trial registration number, digital object identifier (DOI), publication journal, funding sources and conflict of interest.

#### Methodology

\# of participating centers, study setting, blinding methods, phase of study, enrollment duration, randomization and allocation methods, technique for BMD measurement, technique for fracture detection, BTM detection methods and assay types, bone pain scale.

#### Baseline Data

\# randomized, # analyzed, # lost to follow-up, mean age, sex, # postmenopausal, mean duration since diabetes diagnosis, fracture (vertebral and nonvertebral) prevalence at baseline, baseline BMD, BTMs, pain scale measurements.

#### Outcomes

Final BMD measurements or percentage/absolute change in BMD from baseline, # vertebral fracture incidences at latest follow-up, # non-vertebral fracture incidences at latest follow-up. Percentage change in BTMs from baseline, absolute change in pain score from baseline.

#### Other Data

Adverse events, description of anti-diabetic and antiresorptive therapy (i.e. dosage, duration), mean follow-up.

### Statistical Analysis

#### Network Meta-Analysis

We will conduct all statistical analyses using R 3.5.1[38]. We will perform NMAs using the gemtc 0.8-3 library which is based on the Bayesian probability framework[39]. Because we expect significant heterogeneity among studies due to differences in methodology, we will use a random effects model[40].

For Part I of the analysis, we will use patients receiving no active anti-diabetics medication, such as patients managing T2DM using lifestyle choices, as a reference for comparison. If this treatment arm does not exist, placebo or insulin-only patients will be used instead.

For Part II of the analysis, patients receiving no antiresorptive interventions will be used as a reference for comparison. If this treatment arm does not exist, placebo patients will be used instead. To simplify our analysis, we will not take concurrent anti-diabetic medications into account for this portion of the analysis.

For changes in BMD and pain scores, we will report the results of the analysis as weighted mean differences (WMDs) with 95% credible intervals (CrIs) if all included studies utilized the same scale (e.g. if BMD changes are only reported as percentage changes, or if pain outcomes are only reported as 10 point VAS scores). Otherwise, we will report these outcomes as standardized mean differences (SMDs) to include all available RCT data. For BMD outcomes, we will use SMD even if BMD changes can be converted between absolute and percentage changes in order to avoid estimation of the standard deviation (SD) values. However, because SMDs are difficult to interpret for most clinicians, we will supplement our BMD results with weighted mean differences (WMD) as well, considering only percentage changes in BMD[41,42]. BTMs will be analyzed as WMD of percentage changes. Fracture incidences will be reported as odds ratios with corresponding 95% CrIs. We will run all network models for a minimum of 100,000 iterations to ensure convergence.

If there are outcomes for which we did not gather enough information to perform a NMA, we will provide a qualitative description of the available data and study outcomes.

#### Treatment Ranking

We will use the surface under the cumulative ranking curve (SUCRA) scores to provide an estimate as to the ranking of treatments. SUCRA scores range from 0 to 1, with higher SUCRA scores indicating more efficacious treatment arms[43].

#### Missing Data

We will attempt to contact the authors of the original studies to obtain missing or unpublished data. Missing standard deviation values may be imputed using methods described in the Cochrane Handbook for Systematic Reviews of Interventions[44].

#### Heterogeneity Assessment

We will assess statistical heterogeneity within each outcome network using I^2^ statistics and the Cochrane Q test[45]. We will consider an I^2^ index ≥ 50% as an indication for serious heterogeneity, and I^2^ index > 75% as an indication for very serious heterogeneity. We will explore potential sources of heterogeneity using meta-regression analyses.

#### Inconsistency

We will assess inconsistency using the node-splitting method[46]. We will explore any indications of significant inconsistency using meta-regression analyses.

#### Publication Bias

To assess small-study effects within the networks, we will use a comparison-adjusted funnel plot[47]. We will use Egger’s regression test to check for asymmetry within the funnel plot to identify possible publication bias[48].

#### Quality of Evidence

We will use the Confidence in Network Meta-Analysis (CINeMA) web application to evaluate confidence in the findings from our NMA[49]. CINeMA adheres to the GRADE approach for evaluating the quality of evidence by assessing network quality based on six criteria: within-study bias, across-study bias, indirectness, imprecision, heterogeneity and incoherence[50,51].

CINeMA utilizes a frequentist approach to NMAs, which is different from the Bayesian approach used by gemtc. However, previous study has shown that there are no significant differences between frequentist and Bayesian network estimates, therefore the results of the CINeMA analysis should be applicable to our Bayesian networks[52]. We will report the results of our GRADE analysis using a summary of findings table.

#### Meta-Regression

There are several potential factors for increased bone resorption and increased fracture incidences apart from T2DM, such as gender, post-menopausal status, and age[53]. Previous fractures at baseline are also associated with a higher risk of subsequent fractures[54,55]. Variations in these characteristics between studies can result in significant heterogeneity and inconsistency. Therefore, we will conduct meta-regression analyses to check for covariate effects associated with these characteristics.

We will conduct meta-regressions on % female in the patient population, % postmenopausal in the patient population and the median age of the population for BMD, BTM, pain and fracture outcomes. We will also conduct meta-regression on common clinical parameters such as time since diagnosis, duration of drug administration and duration of follow-up for all outcomes. For fracture incidences, we will run a meta-regression on fracture prevalence at baseline. We hypothesize that an increase in mean age, as well as the percentage of females and postmenopausal patients in the population will result in less positive BMD changes, decreased bone formation BTM levels, greater pain and increased fracture incidence.

Longer time since diagnosis will also cause these effects. Similarly, an increase in the number of prevalent fractures at baseline will result in increased fracture incidence. We hypothesize that increased drug duration will increase BMD and bone formation BTM levels, while decreasing pain and fractures. Increased follow-up duration and time since diagnosis will have the opposite effects.

Since we will not consider the effect of concurrent anti-diabetic medications in Part II of our analysis, we will conduct a categorical meta-regression of concurrent anti-diabetic medications for Part II to examine the impact of anti-diabetics. We will also conduct a categorical meta-regression on the location of the studies for both parts of the analysis to examine the impact of differences in the Chinese and Western healthcare environments.

### Patient and Public Involvement

We invited select physicians who are specialized in diabetes and endocrinology or orthopaedics to help us refine our research question as well as primary and secondary outcomes. However, they were not involved in designing any other aspects of this study, nor were they involved in the drafting of this protocol. Due to the nature of our proposed study design, it was not appropriate for us to involve patients in our protocol or study.

## DISCUSSION

Previous NMAs regarding anti-diabetic medications and fracture risks focused on SGLT2 inhibitors and the literature searches were limited to Western databases[25,26]. The Chinese meta-analysis concerning the use of antiresorptive therapies in T2DM patients was limited to alendronate, and only performed searches on Chinese databases[27]. As a result, these latest analyses did not include all available RCT data.

This two-part study aims to significantly expand upon all of the previous analyses by incorporating the entirety of global RCT evidence available. To our knowledge, our proposed study will be the first review to evaluate the relative effects of multiple antiresorptive agents among T2DM patients using a NMA approach, and it will be the most comprehensive analysis evaluating the effect of anti-diabetics on bone health with multi-language search strategies.

Our review will have several strengths. First, we will extend our database search to Chinese databases for Part I of our analysis. Because of China’s immense patient population and regulations that promote pharmaceutical research, the inclusion of Chinese RCTs will help strengthen the power and precision of our analyses[56]. Furthermore, we will use NMA techniques to analyze RCTs concerning antiresorptive pharmacotherapies. This strategy will allow us to include all available treatment arms, including risedronate, zoledronic acid, and calcitonin. We have identified trials examining these treatments, however they were not included in the latest analysis due to limitations with the pairwise meta-analytic study design[23,24,57]. Lastly, we will only include RCT data, and we will use tools such as The Cochrane Collaboration’s tool for assessing risk of bias in randomized trials, CINeMA, and comparison-adjusted funnel plots to evaluate the quality of our included studies and networks.

Our review will also have limitations. Chinese clinicians may not adopt the same procedures and practices as Western clinicians (such as higher drug dosages and different drug formulations); as a result, outcomes from Chinese RCTs may not be applicable to the Western healthcare system. Additionally, we cannot directly compare the efficacy of antiresorptive therapy to anti-diabetics, nor to combinations of antiresorptive therapies and anti-diabetics with our study design.

Despite these limitations, our two-part NMA will likely be the largest quantitative synthesis assessing anti-diabetic and antiresorptive therapies among T2DM patients to date. Our study should help physicians and patients with selecting anti-diabetic regimens that are the most beneficial for T2DM patients’ bone health, as well as selecting the optimal antiresorptive regimen as a concurrent, supplemental therapy. Our study may also highlight promising treatment strategies that were not discussed in the previous analyses, providing physicians and researchers with future research directions.

## Data Availability

Relevant data available upon request.

## ETHICS AND DISSEMINATION

The study will not require ethics approval.

We do not wish to engage in the practice of publishing minimum publishable units (publons)[58]. Therefore, we will attempt to combine the proposed two-part study into a single publication for dissemination, as both parts are highly relevant to the topic of T2DM induced bone disease. However, should the combined publication exceed the word and figure limits imposed by publishers, we will publish the proposed study as two separate publications. The findings of the proposed review will be disseminated in peer-reviewed journals and presented at conferences.

## ACKNOWLEDGEMENTS

We would like to offer our special thanks to Emma Huang, Faculty of Health Sciences, McMaster University for dedicating her time to thoroughly review our protocol.

## AUTHOR STATEMENT

JWD made significant contributions to conception and design of the work, drafted the work, and substantially reviewed it. UA and SD drafted the work, and substantially reviewed it. OC and SS made significant contributions to the methodology of the work. AB made contributions to the conception of the work, substantially reviewed it, and made revisions to the final work.

